# Interim Analysis of Risk Factors for Severe Outcomes among a Cohort of Hospitalized Adults Identified through the U.S. Coronavirus Disease 2019 (COVID-19)-Associated Hospitalization Surveillance Network (COVID-NET)

**DOI:** 10.1101/2020.05.18.20103390

**Authors:** L Kim, S Garg, A O’Halloran, M Whitaker, H Pham, EJ Anderson, I Armistead, NM Bennett, L Billing, K Como-Sabetti, M Hill, S Kim, ML Monroe, A Muse, A Reingold, W Schaffner, M Sutton, HK Talbot, SM Torres, K Yousey-Hindes, R Holstein, C Cummings, L Brammer, A Hall, A Fry, GE Langley

**Author notes:** These authors contributed equally to this work (L Kim, S Garg). Corresponding author: Lindsay Kim, 1600 Clifton Road, MS H24-5, Atlanta, GA 30329; Shikha Garg, 1600 Clifton Road, MS H24-7, Atlanta, GA 30329.

## Abstract

**Background:** As of May 15, 2020, the United States has reported the greatest number of coronavirus disease 2019 (COVID-19) cases and deaths globally.

**Objective:** To describe risk factors for severe outcomes among adults hospitalized with COVID-19.

**Design:** Cohort study of patients identified through the Coronavirus Disease 2019-Associated Hospitalization Surveillance Network.

**Setting:** 154 acute care hospitals in 74 counties in 13 states.

**Patients:** 2491 patients hospitalized with laboratory-confirmed COVID-19 during March 1-May 2, 2020.

**Measurements:** Age, sex, race/ethnicity, and underlying medical conditions.

**Results:** Ninety-two percent of patients had ≥1 underlying condition; 32% required intensive care unit (ICU) admission; 19% invasive mechanical ventilation; 15% vasopressors; and 17% died during hospitalization. Independent factors associated with ICU admission included ages 50-64, 65-74, 75-84 and ≥85 years versus 18-39 years (adjusted risk ratio (aRR) 1.53, 1.65, 1.84 and 1.43, respectively); male sex (aRR 1.34); obesity (aRR 1.31); immunosuppression (aRR 1.29); and diabetes (aRR 1.13). Independent factors associated with in-hospital mortality included ages 50-64, 65-74, 75-84 and ≥85 years versus 18-39 years (aRR 3.11, 5.77, 7.67 and 10.98, respectively); male sex (aRR 1.30); immunosuppression (aRR 1.39); renal disease (aRR 1.33); chronic lung disease (aRR 1.31); cardiovascular disease (aRR 1.28); neurologic disorders (aRR 1.25); and diabetes (aRR 1.19). Race/ethnicity was not associated with either ICU admission or death.

**Limitation:** Data were limited to patients who were discharged or died in-hospital and had complete chart abstractions; patients who were still hospitalized or did not have accessible medical records were excluded.

**Conclusion:** In-hospital mortality for COVID-19 increased markedly with increasing age. These data help to characterize persons at highest risk for severe COVID-19-associated outcomes and define target groups for prevention and treatment strategies.

**Funding Source:** This work was supported by grant CK17-1701 from the Centers of Disease Control and Prevention through an Emerging Infections Program cooperative agreement and by Cooperative Agreement Number NU38OT000297-02-00 awarded to the Council of State and Territorial Epidemiologists from the Centers for Disease Control and Prevention.

## Introduction

In December 2019, an outbreak of a novel coronavirus disease, termed coronavirus disease-2019 (COVID-19), was reported in China caused by a newly identified coronavirus, severe acute respiratory syndrome coronavirus-2 (SARS-CoV-2). Since then, approximately 4.5 million cases of COVID-19 have been reported globally (1). As of May 15, approximately 1.4 million cases, including nearly 86,000 deaths, have been reported in the United States, and case counts continue to rise (1) with evidence of widespread community transmission (2).

Previous reports from China, Italy, and New York City have demonstrated that hospitalized patients are generally older and have underlying medical conditions, such as hypertension and diabetes (3–5). These studies have also found that older patients and those with certain underlying medical conditions like diabetes were at higher risk for severe outcomes (3, 6, 7). Among cases reported to the U.S. Centers for Disease Control and Prevention (CDC) from local and state health departments, the prevalence of underlying medical conditions increased as severity of infections increased (8, 9), although findings were limited by missing or incomplete information. Questions remain about the independent association of sex, race/ethnicity and specific underlying conditions with severe outcomes among persons hospitalized with COVID-19, after adjusting for age and other important potential confounders.

Comprehensive data on U.S. patients with severe COVID-19 infections are needed to better inform clinicians’ understanding of groups at risk for poor outcomes and to inform current prevention efforts and future interventions. We rapidly implemented population-based surveillance for laboratory-confirmed COVID-19-associated hospitalizations, collecting clinical data from hospitalized patients in 154 hospitals in 13 states since March 1, 2020. In this interim analysis restricted to patients who were discharged or died in-hospital and had completed medical chart abstractions, we describe the characteristics of U.S. adults hospitalized with COVID-19 and assess risk factors for intensive care unit (ICU) admission and in-hospital mortality.

## METHODS

### Surveillance Overview

The Coronavirus Disease 2019-Associated Hospitalization Surveillance Network (COVID-NET) has been previously described (10). Eligible COVID-19-associated hospitalizations occurred among persons who (1) resided in a pre-defined surveillance catchment area; and (2) had a positive SARS-CoV-2 test within 14 days prior to or during hospitalization. Hospitalization was defined as admission to an inpatient ward for any length of time, an observation unit stay for ≥24 hours, or a combined stay in an emergency department and observation unit for ≥24 hours.

COVID-NET surveillance occurs in acute care hospitals within 99 counties located in 14 states (California, Colorado, Connecticut, Georgia, Iowa, Maryland, Michigan, Minnesota, New Mexico, New York, Ohio, Oregon, Tennessee, and Utah), covering a catchment population of approximately 32 million persons (∼10% of the U.S. population). Although COVID-NET includes all age groups, for this analysis, we excluded children <18 years of age due to small counts (n=101) and one surveillance site (Iowa) for which medical chart abstractions were not conducted on identified cases. We also excluded patients who were still hospitalized at the time of this analysis and all patients for whom medical chart abstractions had not yet been completed. Because the COVID-19 pandemic limited the ability of surveillance officers to access medical records at facilities, patients were more likely to be included in this analysis if they were hospitalized at facilities that provided surveillance officers remote chart access, participated in Health Information Exchanges, or were able to mail or fax records. COVID-NET surveillance was initiated for cases with hospital admission on or after March 1, 2020.

Laboratory-confirmed COVID-19-associated hospitalizations were identified using laboratory and reportable condition databases, hospital infection control databases, electronic medical records, and/or review of hospital discharge records. Laboratory tests were ordered at the discretion of the treating healthcare provider.

Medical chart reviews for demographic and clinical data were conducted by trained surveillance officers using a standard case report form. Underlying medical conditions were categorized into major groups (Appendix Table 1). Obesity and severe obesity were defined as a calculated body mass index (BMI) ≥30 kg/m^2^ and BMI ≥40 kg/m^2^, respectively. Chest radiograph results were obtained from the radiology reports and not from review of the original radiograph. We defined severe outcomes as either ICU admission or in-hospital mortality. We hypothesized that increasing age and underlying medical conditions would be associated with an increased risk of ICU admission and in-hospital mortality.

**Table 1.** Demographic and clinical characteristics of adults hospitalized with COVID-19 — COVID-NET, 13 sites (N=2,491)

|  | n, % |
| --- | --- |
| **Age in years (median, IQR)** | 62, 50–75 |
| **Age category** |  |
| 18–39 | 302 (12.1) |
| 40–49 | 319 (12.8) |
| 50–64 | 744 (29.9) |
| 65–74 | 478 (19.2) |
| 75–84 | 397 (15.9) |
| 85+ | 251 (10.1) |
| **Male** | 1,326 (53.2) |
| **Race/ethnicity (n=2,490)** |  |
| Non-Hispanic White | 1,178 (47.3) |
| Non-Hispanic Black | 755 (30.3) |
| Hispanic | 306 (12.3) |
| Non-Hispanic Other* | 158 (6.3) |
| Unknown | 93 (3.7) |
| **Residence at time of hospitalization (n=2,482)** |  |
| Private residence | 1,899 (76.5) |
| Facility† | 495 (19.9) |
| Homeless/Shelter | 40 (1.6) |
| Other‡ | 45 (1.8) |
| Unknown | 3 (0.1) |
| **Smoker (n=2,489)** |  |
| Current | 150 (6.0) |
| Former | 642 (25.8) |
| No or Unknown | 1,697 (68.2) |
| **Any underlying condition§ (n=2,489)** | 2,278 (91.5) |
| Hypertension (n=2,488) | 1,428 (57.4) |
| Obesity¶ (n=2,332) | 1,154 (49.7) |
| Severe obesity¶ (n=2,332) | 325 (14.0) |
| Chronic metabolic disease (n=2,486) | 1,024 (41.2) |
| Diabetes mellitus (n=2,486) | 819 (32.9) |
| Chronic lung disease (n=2,484) | 747 (30.1) |
| Asthma (n=2,484) | 314 (12.6) |
| Chronic Obstructive Pulmonary Disease (n=2,484) | 266 (10.7) |
| Cardiovascular disease (n=2,486) | 859 (34.6) |
| Coronary artery disease (n=2,486) ¶ | 352 (14.2) |
| Congestive heart failure (n=2,486) | 284 (11.4) |
| Neurologic disease (n=2,484) | 548 (22.1) |
| Renal disease (n=2,488) | 386 (15.5) |
| Immunosuppressive condition (n=2,487) | 263 (10.6) |
| Gastrointestinal or Liver disease (n=2,486) | 118 (4.7) |
| Hematologic condition (n=2,483) | 80 (3.2) |
| Rheumatologic or autoimmune disease (n=2,486) | 77 (3.1) |
| Pregnancy (n=279)** | 36 (12.9) |
| **Number of underlying medical conditions**<br>**(by major category) (n=2,490)** |  |
| 0 | 212 (8.5) |
| 1 | 480 (19.3) |
| 2 | 510 (20.5) |
| 3+ | 1,288 (51.7) |
| **Outpatient use of medications** |  |
| ACE-inhibitor (n=1,892) | 316 (16.7) |
| Angiotensin receptor blockers (ARBs) (n=1,895) | 257 (13.6) |

IQR = interquartile range; ACE = angiotensin-converting enzyme inhibitors
\*Non-Hispanic Other includes: Non-Hispanic American Indian or Alaskan Native (n=24), Non-Hispanic Asian (n=128), and Non-Hispanic multiracial (n=6)
\*Facility includes rehabilitation facilities, assisted living/residential care, group homes, nursing homes, skilled nursing facilities, long-term care facilities, long-term acute care hospitals, alcohol/drug treatment centers, and psychiatric facilities.
\*Other includes home with services (n=43), correctional facility (n=1), and hospice (n=1).
§See Supplementary Table 1 for definitions of underlying medical conditions.
||Obesity is defined as body mass index (BMI) $\geq 30$ kg/m<sup>2</sup>, and severe obesity is defined as BMI $\geq 40$ kg/m<sup>2</sup>.
¶Includes coronary artery disease, history of coronary artery bypass grafting, and history of myocardial infarction
\*\*Denominator includes women aged 15–49 years.

### Statistical Analysis

After the exclusions noted above, we included adults hospitalized within 154 acute care hospitals in 74 counties in 13 states with an admission date during March 1–May 2, 2020 who had either been discharged from the hospital or died during hospitalization and had complete medical chart abstractions. We calculated proportions using the number of patients with data available on each characteristic as the denominator.

To construct multivariable models for ICU admission and in-hospital mortality, we first assessed collinearity among underlying medical condition categories and outpatient use of ACE-inhibitors and ARBs. We examined the association of demographic factors, underlying medical conditions, and outpatient use of ACE-inhibitors and ARBs with ICU admission and in-hospital death using chi square tests. Variables considered for inclusion in the final models included current or former smoker, hypertension, obesity, diabetes, chronic lung disease (CLD), cardiovascular disease (CVD) (excluding hypertension), neurologic disorders, renal disease, immunosuppression, gastrointestinal/liver disease, hematologic conditions, rheumatologic/autoimmune conditions, and outpatient use of ACE-inhibitors or ARBs. All multivariable models included age categorized into the following groups (18–39, 40–49, 50– 64, 65–74, 75–84, and ≥85 years), sex, and race/ethnicity. Other variables with p-values <0.10 in bivariate analyses were included in the multivariable analyses. Log-linked Poisson generalized estimating equations regression with an exchangeable correlation matrix (11, 12), clustered by site, was used to generate adjusted risk ratios (aRR), 95% confidence intervals (CI), and two-sided p-values for the risk of ICU admission and in-hospital death. We also constructed separate multivariable models to examine the association between the number of underlying medical conditions and ICU admission or in-hospital death. Two-sided p-values <0.05 were considered statistically significant. All analyses were performed using the SAS 9.4 software (SAS Institute Inc., Cary, NC, USA).

These data were collected as part of routine public health surveillance and determined to be non-research by CDC. Participating sites obtained approval for the COVID-NET surveillance protocol from their respective state and local IRBs, as required.

## RESULTS

A total of 16,318 hospitalized COVID-19 patients were reported to COVID-NET with an admission date during March 1–May 2, 2020. After excluding 101 pediatric patients, 74 patients from Iowa where detailed medical chart abstractions are not conducted, and 13,652 patients who were either still hospitalized or did not yet have completed medical chart abstractions, 2,491 (15%) COVID-19-associated hospitalized adults from 13 surveillance sites were included in this analysis (Figure 1). Patients came estimating equations with an exchangeable correlation matrix (11, 12), clustered by site, was from 43% (154/357) of the acute care hospitals included in COVID-NET surveillance across the 13 surveillance sites (Appendix Table 2). The percentage of facilities contributing data out of the total number of facilities by site ranged from 19% to 100%. The median age of included and excluded patients (62 vs. 63 years, respectively) was similar (Appendix Table 3). The highest proportion of patients included in this analysis were from Minnesota (20%), Tennessee (20%), New York (12%), and Maryland (10%), and Connecticut (9%) (Appendix Table 3).

**Figure 1.**
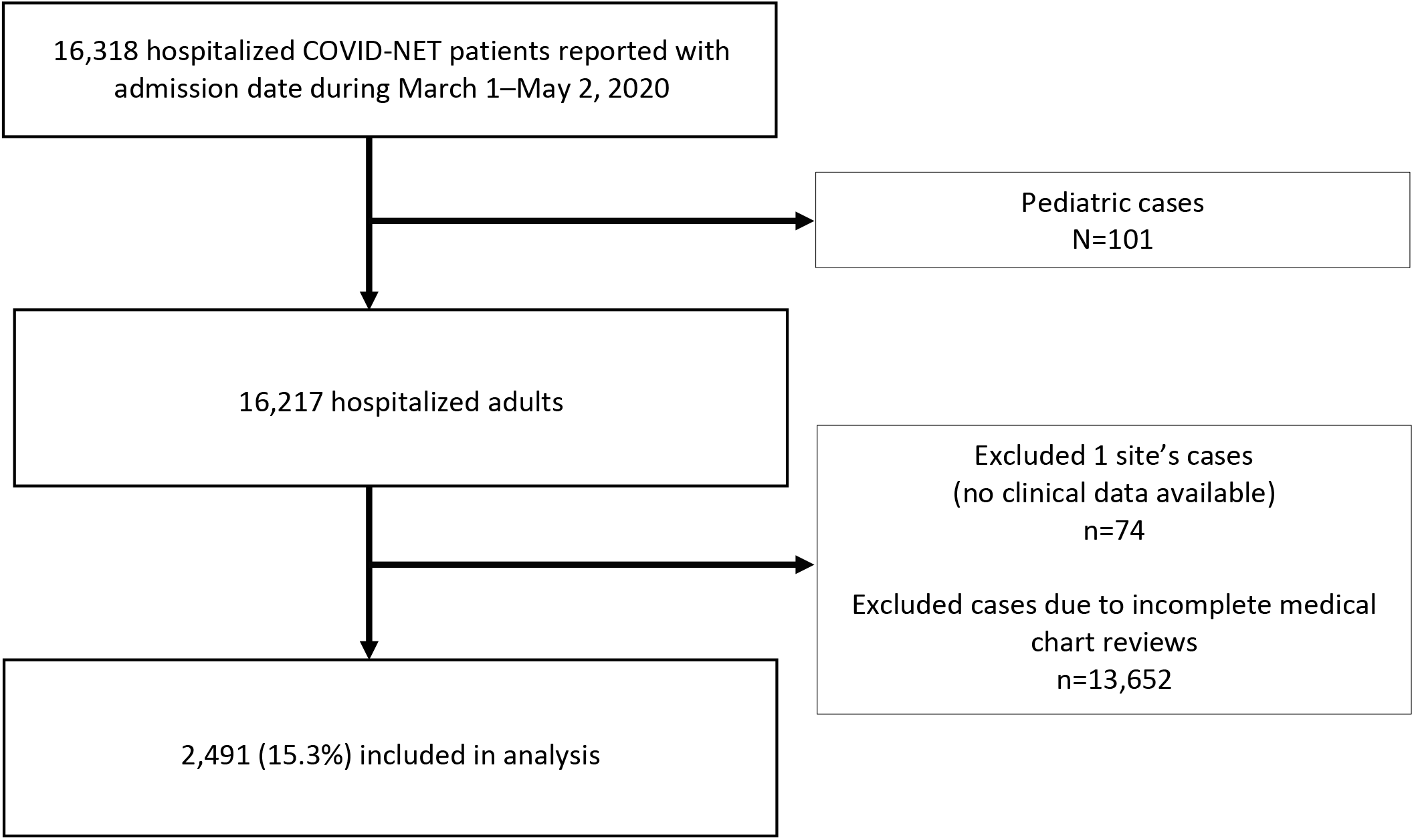
Flow diagram of analytic sample.

### Characteristics of hospitalized patients with COVID-19

Among the 2,491 hospitalized adults, median age was 62 years (interquartile range (IQR), 50– 75), and almost 75% were ≥50 years (Table 1). Forty-seven percent of patients (n=1178/2490) were non-Hispanic whites, 30% non-Hispanic blacks (n=755/2490), and 12% Hispanics (n=306/2490). Nearly one-third of patients were current or former smokers.

Almost all patients (n=2278/2489, 92%) had ≥1 underlying medical condition, with hypertension (n=1428/2488, 57%), obesity (n=1154/2332, 50%), and chronic metabolic disease (n=1024/2486, 41%) most frequently documented. Among patients with chronic metabolic disease, 80% (n=819/1024) had diabetes mellitus; hypertension alone was documented in only 4% (n=91/2490) of patients. Seventeen percent (n=316/1892) took ACE-inhibitors, and 14% (n=257/1895) took ARBs prior to hospitalization. The proportion of patients with any documented underlying medical condition increased with age (p<0.05)(Figure 2A). Prevalence of CLD, neurologic conditions, obesity, and renal disease varied between males and females (p<0.05, Figure 2B). CVD, CLD, and neurologic conditions were more prevalent among non-Hispanic whites, while diabetes, hypertension, obesity and renal disease were more common among non-Hispanic blacks (p<0.05, Figure 2C).

**Figure 2.**
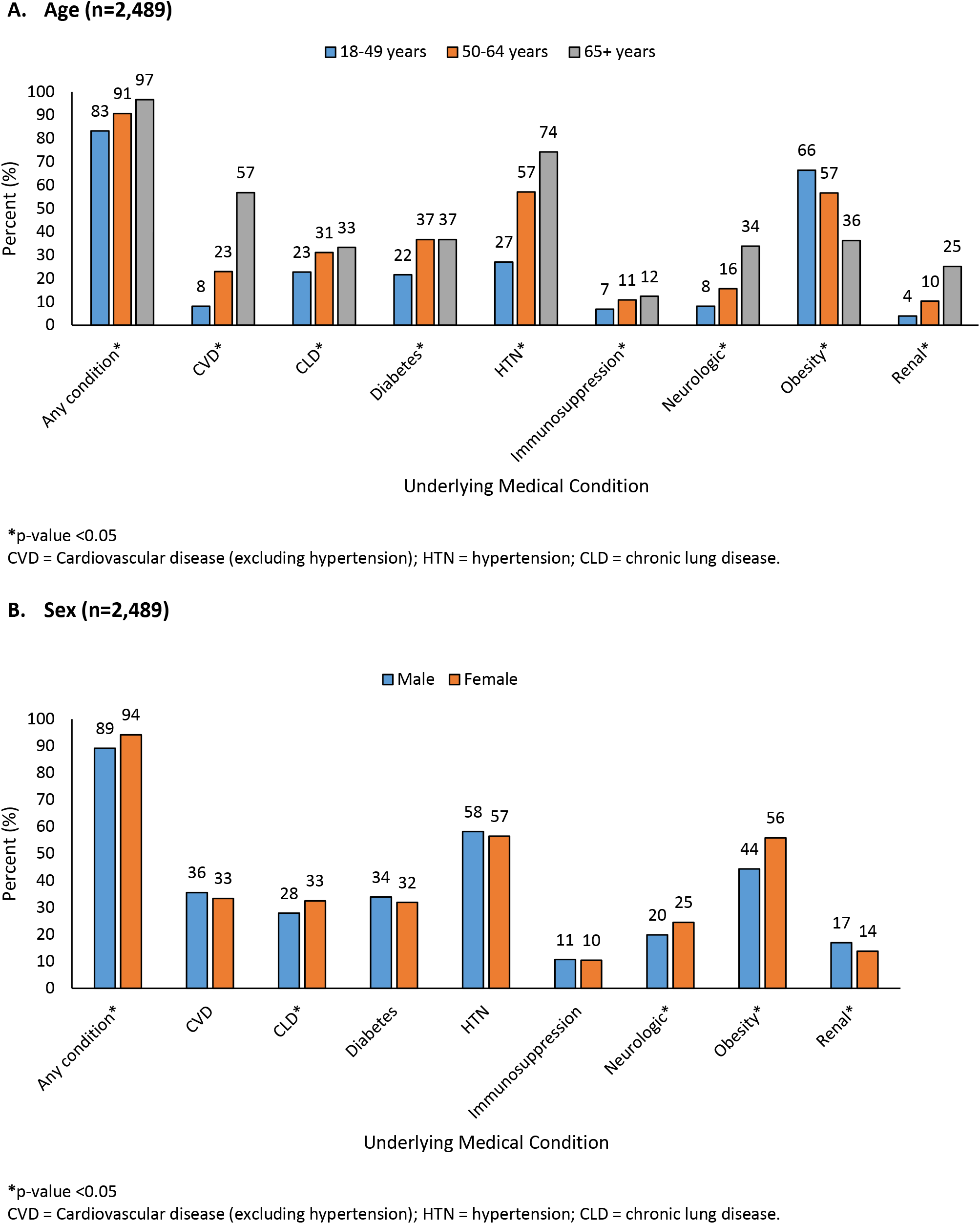
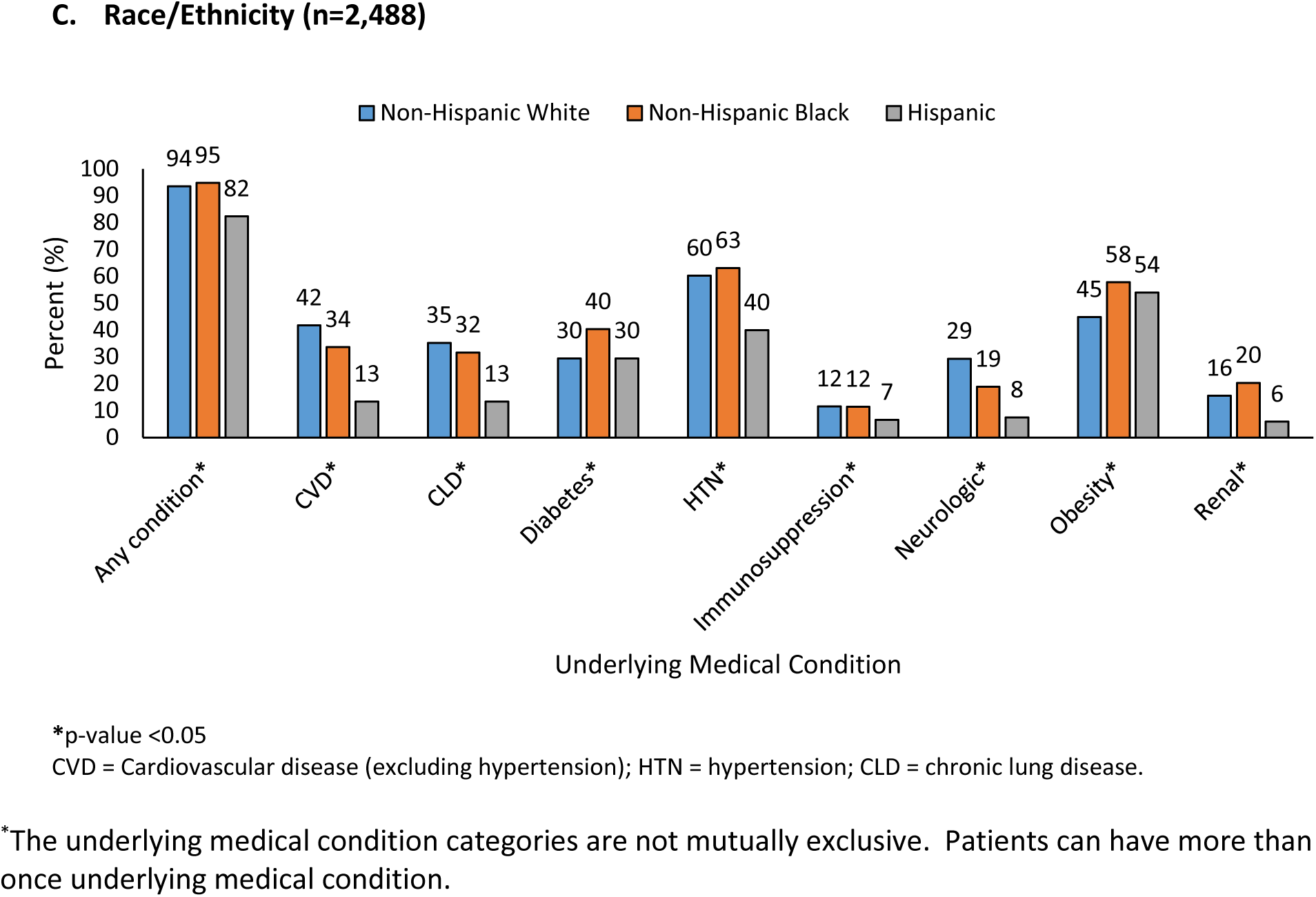
Select underlying medical conditions^*^ of adults hospitalized with COVID-19, by age, sex, and race/ethnicity — COVID-NET, 13 sites (N=2,491)

Cough (75%), fever or chills (74%), and shortness of breath (70%) were commonly documented symptoms at admission (Table 2 and Appendix Table 4). Gastrointestinal symptoms, including nausea, vomiting, and diarrhea, were documented in almost 30% of patients. Median length of hospitalization was 6 days (IQR, 3–11), and median days from symptom onset to hospital admission was 6 days (IQR, 3–8). Median values of initial vital signs were within normal range, except for elevated blood pressure (Table 2). Thirty-three individuals had a pathogen detected from positive blood cultures (Appendix Table 5). Viral co-detections from respiratory specimens were rare among those who were tested (n=38/1549, 2.5%) (Appendix Table 6). Among 1932 patients with chest radiograph performed, 92% (n=1769) were documented as abnormal with infiltrate or consolidation (n=1574/1932, 81%) documented most frequently (Appendix Table 7). Ninety-five percent (n=540/566) of patients with chest computerized tomography (CT) had abnormal findings, and ground glass opacity was documented in 62% (n=350/566) (Appendix Table 7).

**Table 2.** Clinical course, interventions, and outcomes of adults hospitalized with COVID-19 — COVIDNET, 13 sites (N=2,491)

|  | % |
| --- | --- |
| **Symptoms on admission\* (n=2,482)** |  |
| Cough | 1,855 (74.7) |
| Fever or chills | 1,835 (73.9) |
| Shortness of breath | 1,740 (70.1) |
| Muscle aches/myalgias | 722 (29.1) |
| Diarrhea | 676 (27.2) |
| Nausea or vomiting | 621 (25.0) |
| **Hospitalization length of stay in days, median (IQR) (n=2,487)** | 6 (3–11) |
| **Days from symptom onset to hospitalization (median, IQR) (n=1,937)** | 6 (3–8) |
| **Initial vital signs** |  |
| Temperature (°Celsius, median, IQR) (n=2,469) | 37.4 (36.9–38.1) |
| Heart rate (median, IQR) (n=2,479) | 95 (83–108) |
| Systolic blood pressure (median, IQR) (n=2,483) | 132 (118–147) |
| Respiratory rate (median, IQR) (n=2,461) | 20 (18–23) |
| Oxygen saturation (among those on room air) (median, IQR) (n=1,969) | 94 (92–97) |
| **Initial laboratory values** |  |
| White blood cell count (median, IQR) – per mm<sup>3</sup> (n=2,458) | 6.3 (4.7–8.5) |
| Hematocrit (median, IQR) - % (n=2,461) | 40.6 (36.9–43.9) |
| Platelet count (median, IQR) – per mm<sup>3</sup> (n=2,461) | 195.0 (156.0–249.0) |
| Sodium (median, IQR) – mmol/L (n=2,460) | 137.0 (134.0–139.0) |
| Blood Urea Nitrogen (median, IQR) – mg/dl (n=2,443) | 16.0 (11.0–25.0) |
| Creatinine (median, IQR) – mg/dl (n=2,462) | 1.0 (0.8–1.4) |
| Glucose (median, IQR) – mg/dl (n=2,459) | 117.0 (102.0–149.0) |
| Aspartate transaminase (median, IQR) – U/L (n=2,149) | 40.0 (28.0–61.0) |
| Alanine aminotransferase (median, IQR) – U/L (n=2,164) | 31.0 (20.0–50.0) |
| Arterial pH (median, IQR) (n=487) | 7.35 (7.40–7.45) |
| **Abnormal chest X-ray during hospitalization (n=1,932)** | 1,769 (91.6) |
| **Abnormal chest CT during hospitalization (n=566)** | 540 (95.4) |
| Ground glass opacities (n=566) | 350 (61.8) |
| **Investigational medication regimens for COVID-19† (n=2,482)** | 1,125 (45.3) |
| Hydroxychloroquine‡ (n=2,479) | 1,065 (43.0) |
| Azithromycin + ≥ 1 other COVID-19 treatment (n=2,479) | 725 (29.2) |
| Tocilizumab (n=2,479) | 103 (4.2) |
| Atazanavir (n=2,479)§ | 94 (3.8) |
| Remdesivir (n=2,479)† | 53 (2.1) |
| Lopinavir/ritonavir (n=2,479)§ | 27 (1.1) |
| Convalescent plasma (n=2,479) | 9 (0.4) |
| Chloroquine (n=2,479) | 7 (0.3) |
| Sarilumab (n=2,479)† | 6 (0.2) |
| Investigational drug (not specified) RCT (n=2,479) | 1 (0.0) |
| **ICU Admission (n=2,490)** | 798 (32.0) |
| ICU length of stay (days, median, IQR) (n=771) | 6 (2–11) |
| **Highest level of respiratory support required (n=2,489)** |  |
| Invasive mechanical ventilation | 462 (18.6) |
| BIPAP or CPAP | 82 (3.3) |
| High flow nasal cannula | 170 (6.8) |
| **ECMO (n=2,487)** | 9 (0.4) |
| **Vasopressor use (n=2,486)** | 373 (15.0) |
| **Systemic steroids (n=2,489)** | 321 (12.9) |
| **Renal replacement therapy (n=2,487)** | 115 (4.6) |
| **Discharge Diagnoses<sup>II</sup>** |  |
| Pneumonia (n=2,485) | 1,395 (56.1) |
| Acute respiratory failure (n=2,487) | 999 (40.2) |
| Acute renal failure (n=2,485) | 456 (18.4) |
| Sepsis (n=2,479) | 443 (17.9) |
| Acute respiratory distress syndrome (n=2,485) | 255 (10.3) |
| Encephalitis (n=2,482) | 151 (6.1) |
| Congestive heart failure (n=2,485) | 51 (2.1) |
| Asthma exacerbation (n=2,486) | 43 (1.7) |
| COPD exacerbation (n=2,486) | 39 (1.6) |
| Acute myocardial infarction (n=2,485) | 38 (1.5) |
| **In-hospital death (n=2,490)** | 420 (16.9) |

IQR = interquartile range; CT = computed tomography; ICU = intensive care unit; BIPAP = bilevel positive airway pressure; CPAP = continuous positive airway pressure; ECMO = extracorporeal membrane oxygenation; COPD = chronic obstructive pulmonary disease
\*See Supplementary Table 2 for additional symptom data.
\*Not mutually exclusive categories
\*Includes randomized controlled trials where it cannot be determined whether the case received treatment vs. placebo (remdesivir, 24; hydroxychloroquine 15; and sarilumab 5).
§Persons with HIV/AIDS were excluded.
<sup>II</sup>Discharge diagnoses recorded from the hospital discharge summary and not based on ICD-10 discharge codes.

Forty-five percent (n=1125/2482) of patients received investigational medication regimens for COVID-19 during hospitalization (Table 2). The most common regimens included hydroxychloroquine (n=1065/2479, 43%) and the combination of azithromycin and ≥1 COVID-19 treatment (n=725/2479, 29%) (non-mutually exclusive categories). The most frequent discharge diagnoses recorded in hospital discharge summaries were pneumonia (n=1395/2485, 56%), acute respiratory failure (n=999/2487, 40%), acute renal failure (n=456/2,485, 18%), and sepsis (n=443/2,479, 18%).

Thirty-two percent (n=798/2490) of patients required ICU admission, with a median length of ICU stay of 6 days (range, 1–41; IQR, 2–11) (Table 2). Median days from symptom onset to ICU admission was 7 days (range, 0–25; IQR, 4–10), and median days from hospital admission to ICU admission was 1 day (range, 0–19; IQR, 0–2). Among 2,489 hospitalized patients, the highest respiratory support received was invasive mechanical ventilation in 19% (n=462), bilevel positive airway pressure (BIPAP) or continuous positive airway pressure (CPAP) in 3% (n=82), and high flow nasal cannula (HFNC) in 7% (n=170). Fifty-three percent (n=246/462) of patients that received invasive mechanical ventilation died in-hospital (median age, 71 years; IQR, 62–79); the proportion of patients receiving mechanical ventilation who died increased with age (p<0.0001). Vasopressors were used in 15% (n=373/2486) of patients, while renal replacement therapy was used in 5% (n=115/2487). As age increased, so did the proportion of patients who required ICU admission, invasive mechanical ventilation, and vasopressors (p<0.05, Figure 3A). Males were admitted to the ICU and treated with invasive mechanical ventilation, HFNC, or vasopressors more frequently than females (p<0.05) (Figure 3B). Non-Hispanic whites more frequently received BIPAP, CPAP or HFNC (p<0.05, Figure 3C).

**Figure 3.**
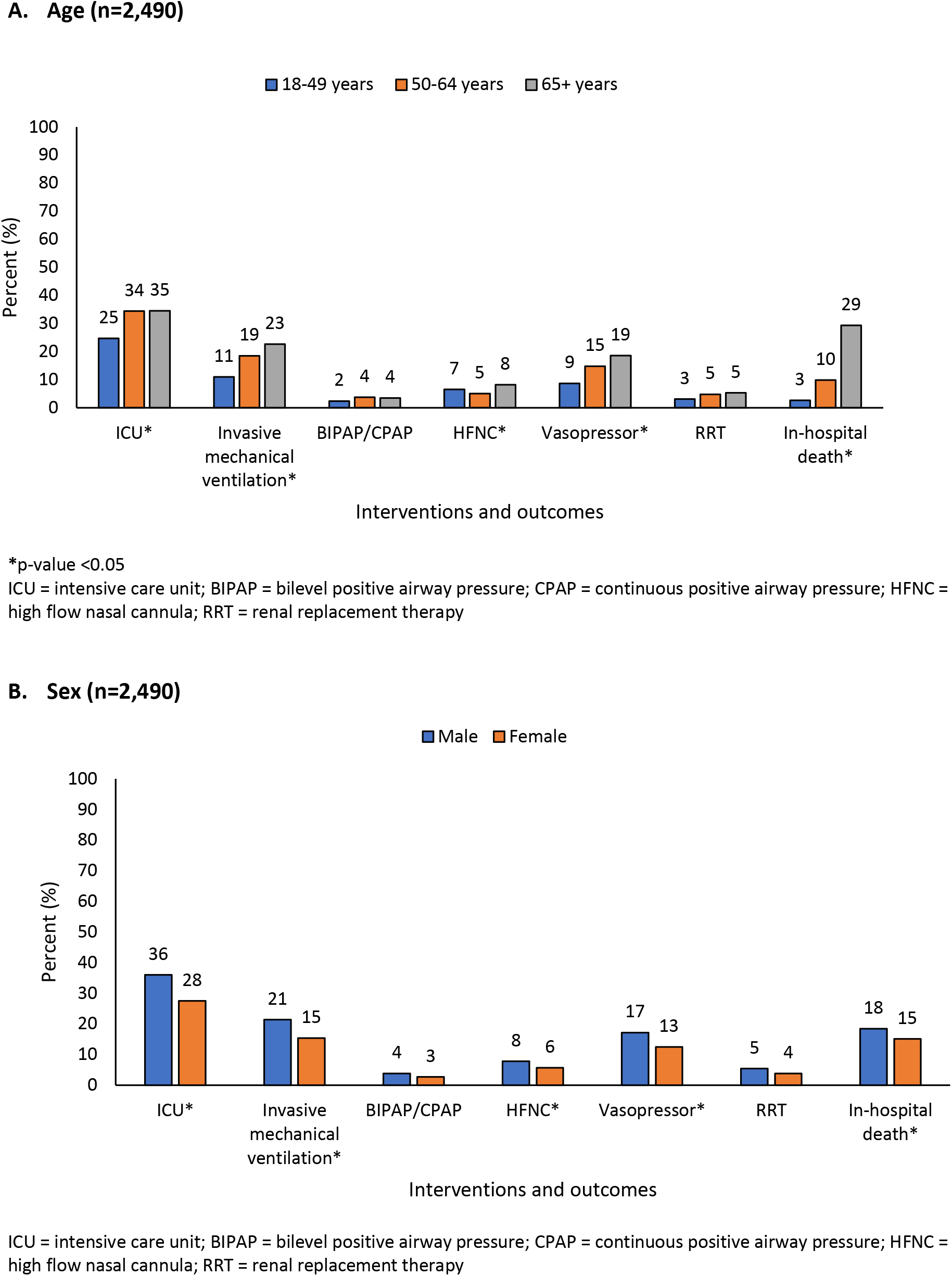
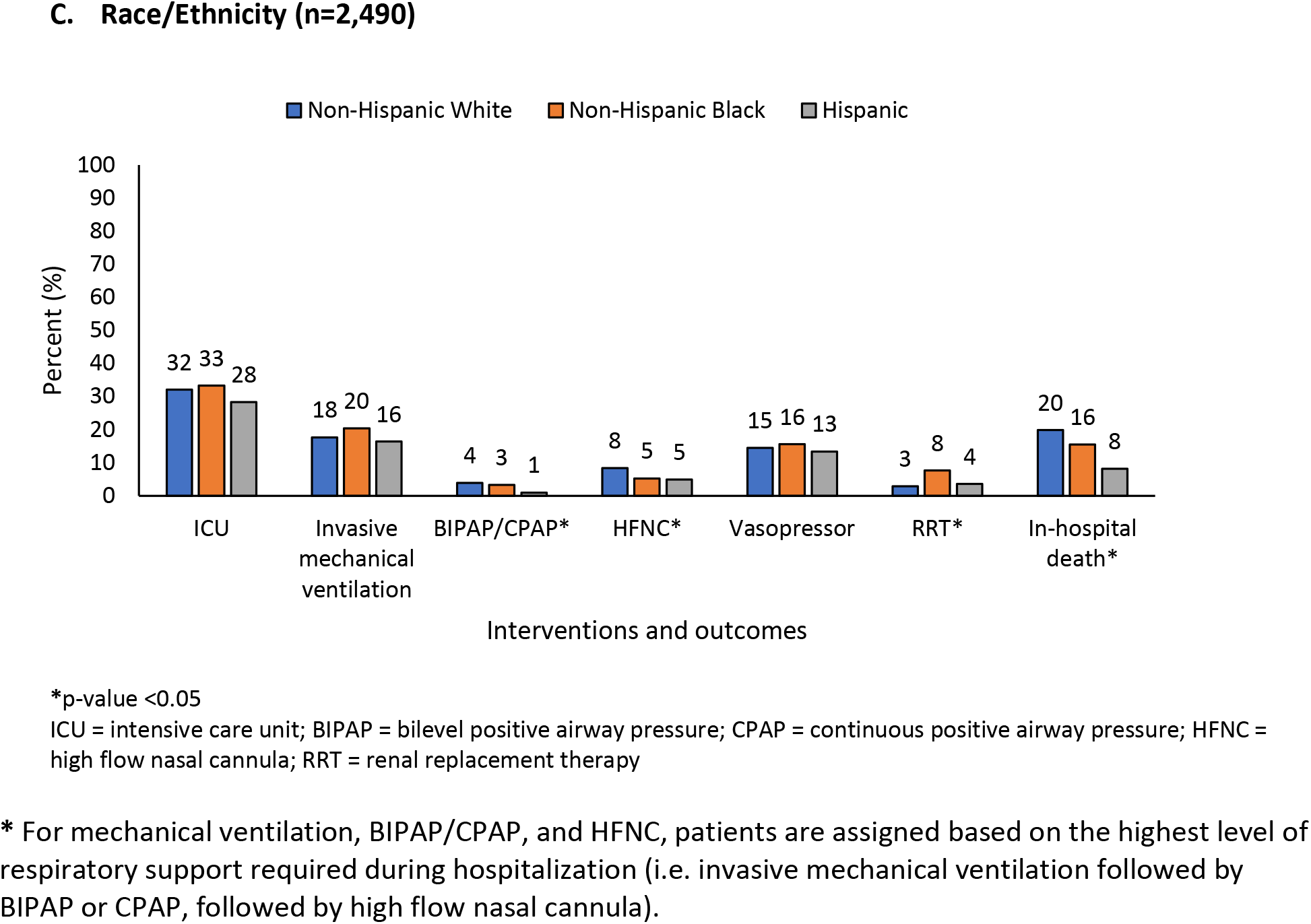
Interventions and outcomes of adults hospitalized with COVID-19, by age, sex, and race/ethnicity — COVID-NET, 13 sites (N = 2,491)

Overall, seventeen percent (n=420/2490) of patients died during hospitalization (Table 2). The proportion of patients who died increased with increasing age groups, ranging from 3% among 18–49 years to 10% among 50–64 years to 29% among ≥65 years (Figure 3A). Males died more frequently compared to females (p<0.05) (Figure 3B), as did non-Hispanic whites compared to other race/ethnicities (p<0.05, Figure 3C).

Among 420 patients who died, median age was 76 years (range, 24–97; IQR, 66–85); 58% (n=244) were male; 71% (n=299) were admitted to the ICU; and 59% (n=246) received invasive mechanical ventilation. The median length of hospitalization among patients who died was 7 days (range, 0–40; IQR, 4–12).

### Risk factors for ICU admission and death

Factors independently associated with ICU admission included age 50–64 years (adjusted risk ratio (aRR) = 1.53; 95% confidence interval (CI), 1.28 to 1.83); 65–74 years (aRR = 1.65; CI, 1.34 to 2.03); 75–84 years (aRR = 1.84; CI, 1.60 to 2.11); ≥85 years (aRR = 1.43; CI, 1.00 to 2.04); male sex (aRR = 1.34; CI, 1.20 to 1.50); obesity (aRR = 1.31; CI, 1.16 to 1.47); diabetes (aRR = 1.13; CI, 1.03 to 1.24); and immunosuppression (aRR = 1.29; CI, 1.13 to 1.47) (Table 3A).

**Table 3.**
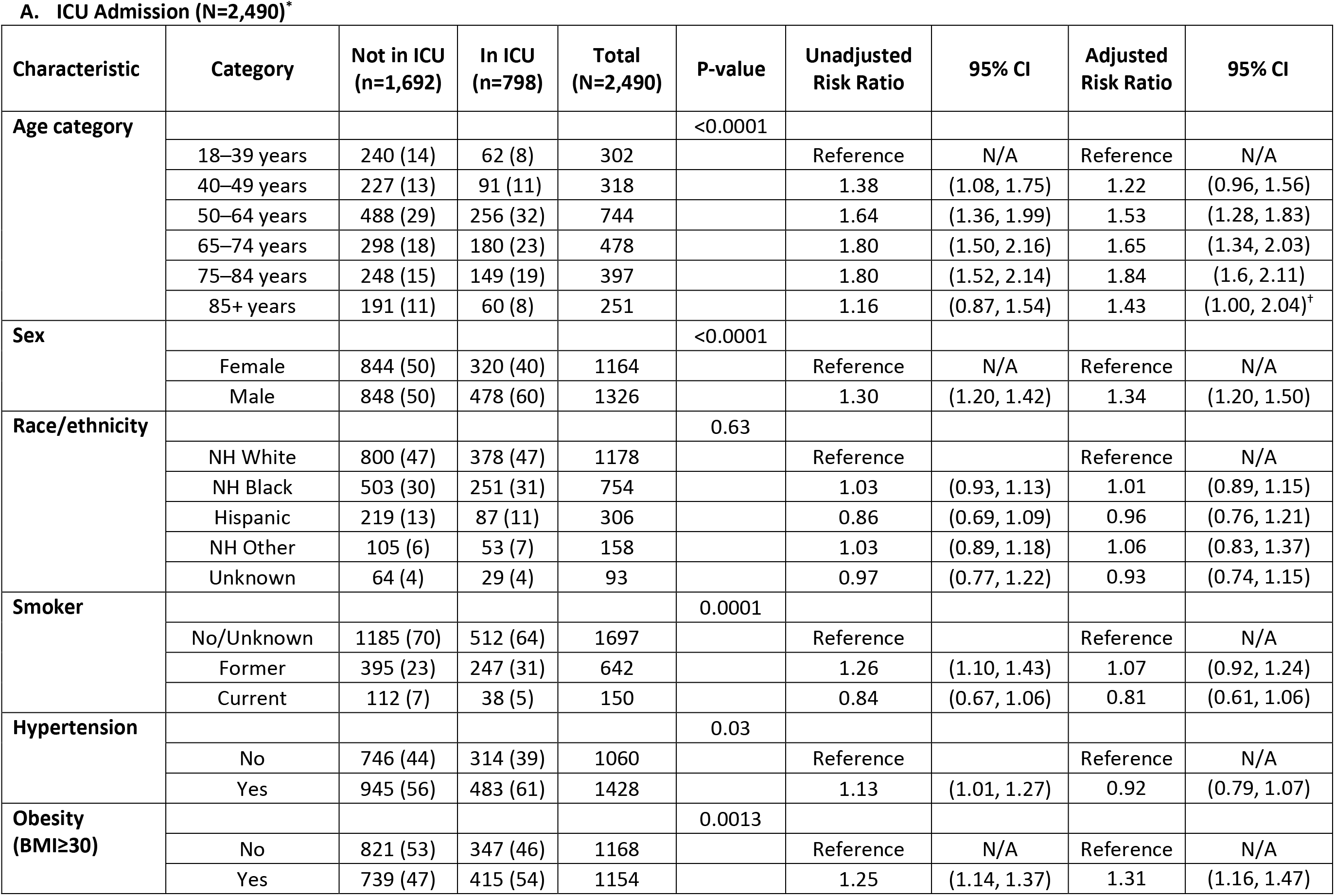
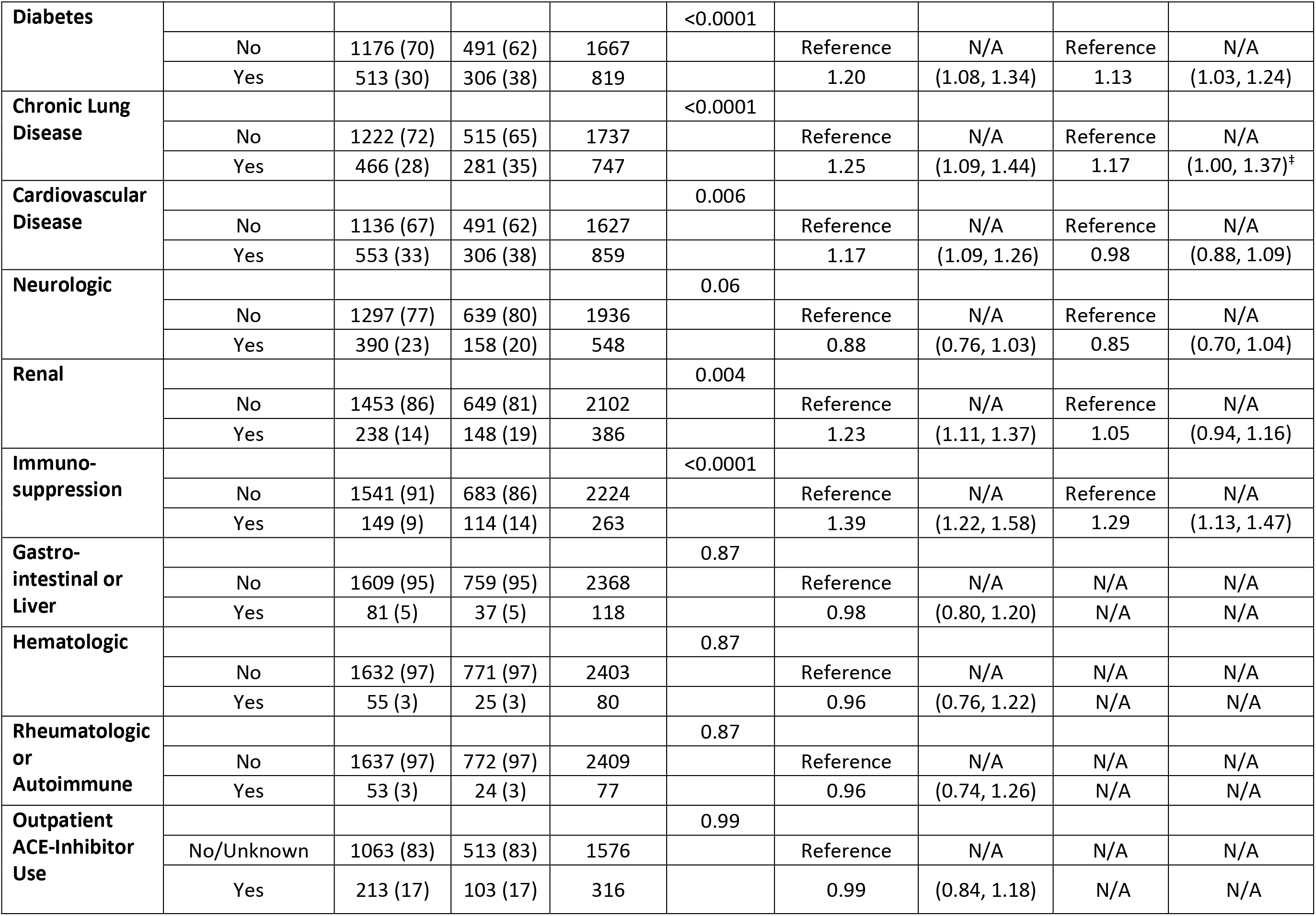
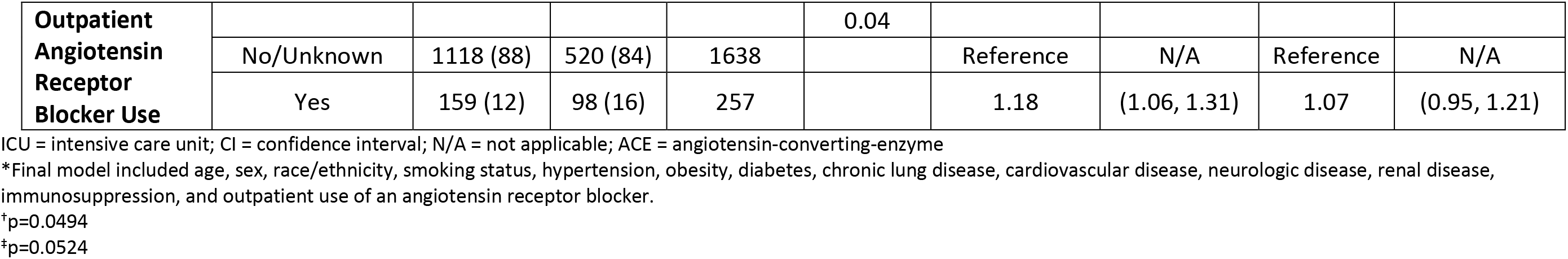
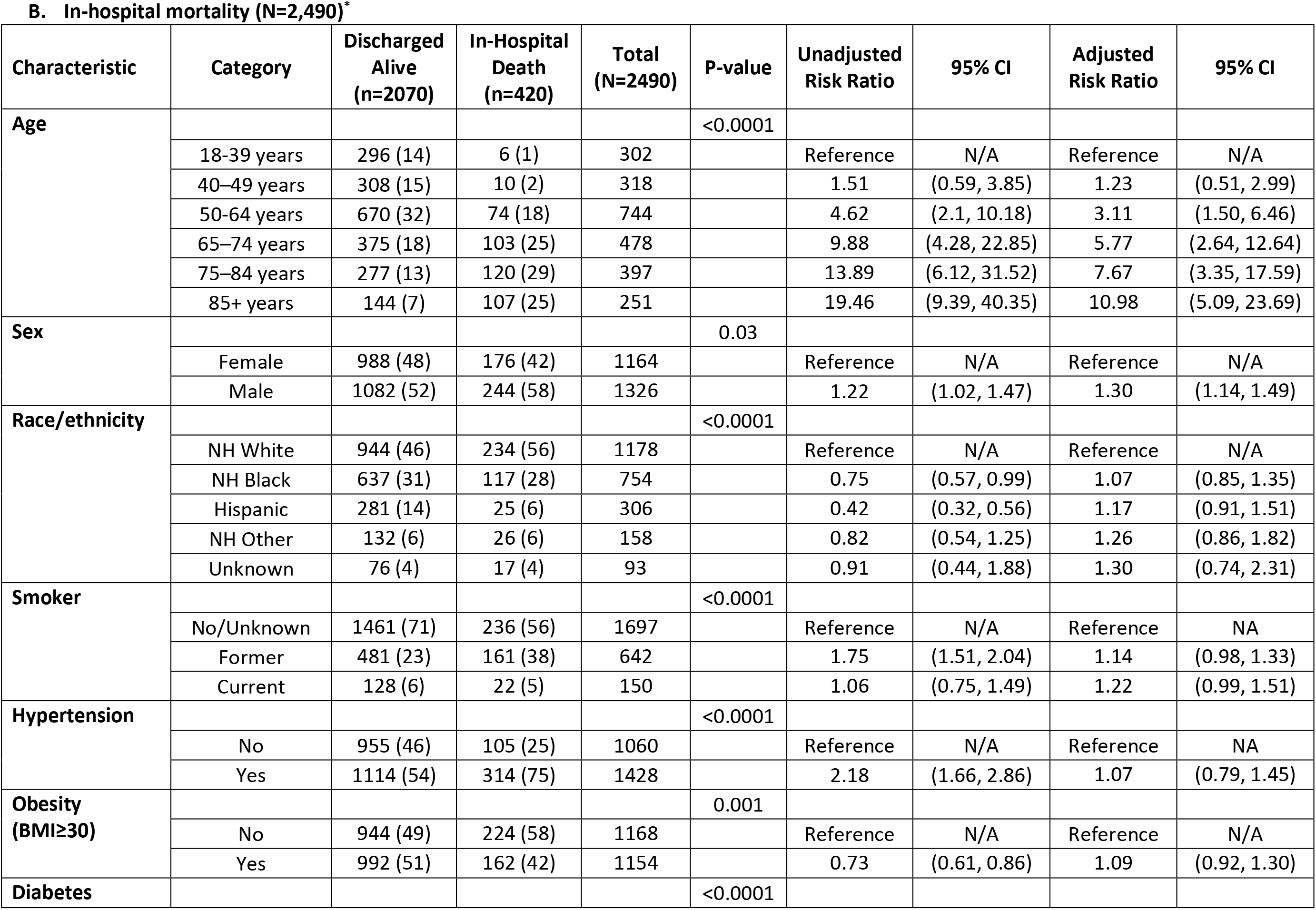
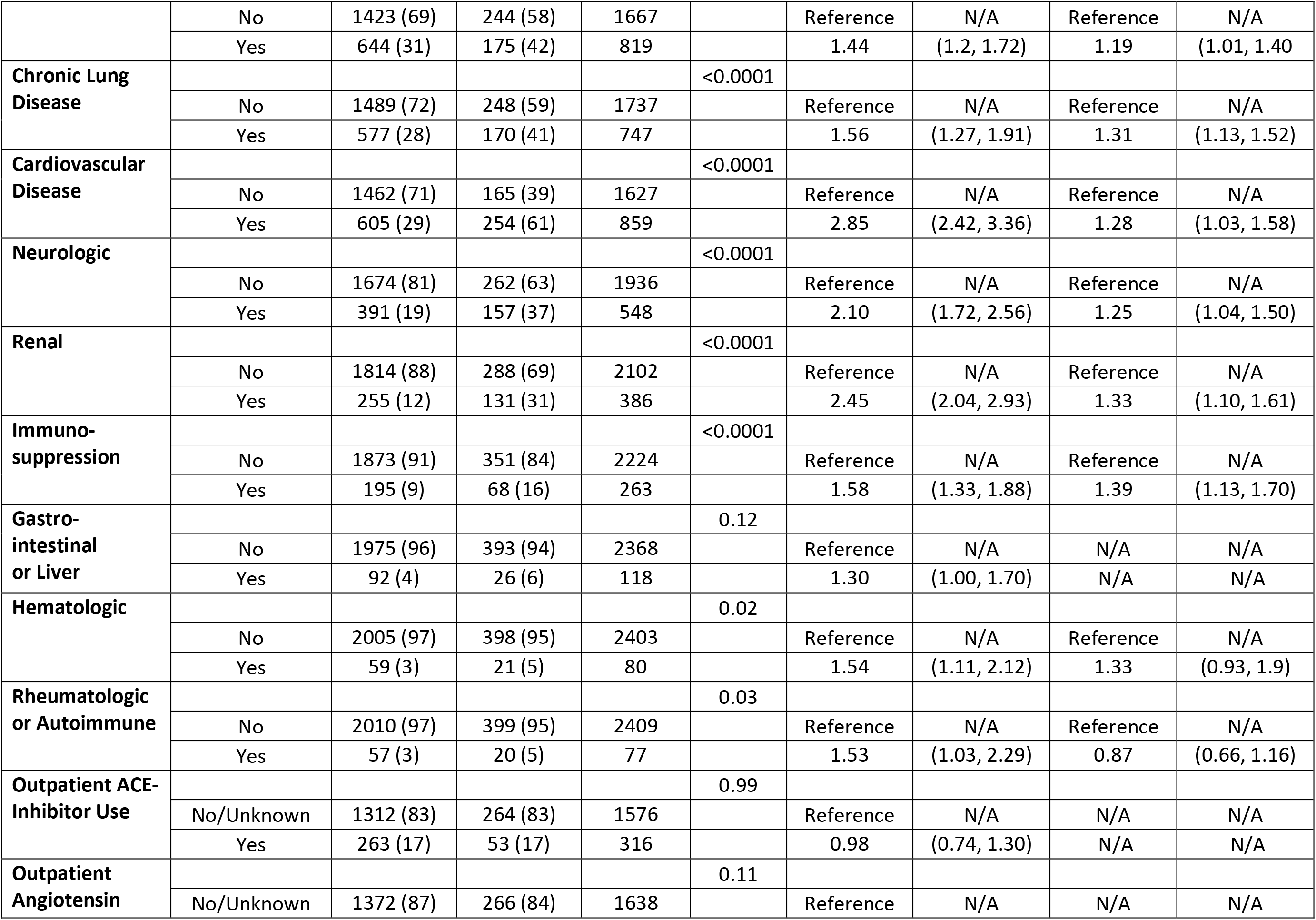
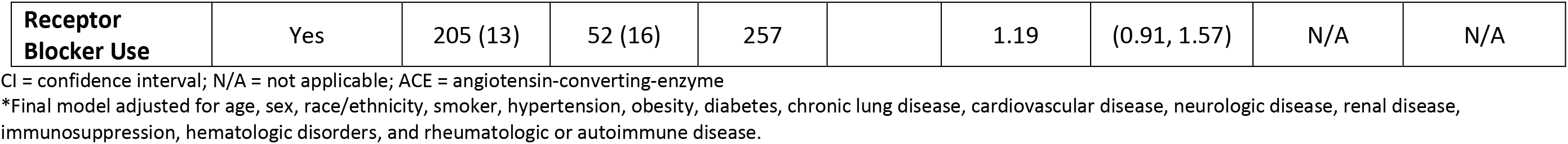
Risk factors for ICU admission and in-hospital mortality — COVID-NET, 13 sites (N=2,491)

Independent factors associated with in-hospital mortality included age 50–64 years (aRR = 3.11; CI 1.50 to 6.46); age 65–74 years (aRR = 5.77; CI, 2.64 to 12.64); age 75–84 years (aRR = 7.67; CI, 3.35 to 17.59); age ≥85 years (aRR = 10.98; CI, 5.09 to 23.69); male sex (aRR = 1.30; CI, 1.14 to 1.49); diabetes (aRR = 1.19; CI, 1.01 to 1.40); CLD (aRR = 1.31; CI, 1.13 to 1.52); CVD (aRR = 1.28; CI, 1.03 to 1.58); neurologic disorders (aRR = 1.25; CI, 1.04 to 1.50); renal disease (aRR = 1.33; CI, 1.10 to 1.61); and immunosuppression (aRR = 1.39; CI, 1.13 to 1.70) (Table 3B).

Having ≥3 underlying medical conditions was significantly associated with higher risk of ICU admission and death after adjusting for age group, sex, and race/ethnicity (Appendix Table 8).

## DISCUSSION

Using a geographically diverse, multi-site, population-based U.S. surveillance system, we found that among adults hospitalized with laboratory-confirmed COVID-19, almost one-third required ICU admission, 19% received invasive mechanical ventilation, and 17% died during hospitalization. About 75% of patients were ≥50 years, and > 90% had underlying medical conditions. Older age, being male, and the presence of certain underlying medical conditions were associated with a higher risk of ICU admission and in-hospital mortality. Race/ethnicity was not independently associated with either outcome among hospitalized patients. This information can alert healthcare providers to patients at greatest risk of severe outcomes and help target prevention strategies and future interventions.

In a published COVID-NET analysis, we found that when comparing the racial/ethnic distribution of residents of the surveillance catchment areas to the racial/ethnic distribution of COVID-19-associated hospitalizations, non-Hispanic blacks were disproportionately hospitalized with COVID-19 compared to non-Hispanic whites (10). In this analysis, however, we found that once hospitalized, non-Hispanic blacks did not have increased risk of poorer outcomes compared to other race/ethnicities after adjusting for age and underlying conditions. In a preprinted article of U.S. Veterans seeking care at VA Hospitals, Rentsch et al. found no association between black race and ICU admission (13). Similarly, a large study of patients hospitalized in New York City did not find race/ethnicity to be associated with ICU admission or death (4).

COVID-19-associated hospitalizations, ICU admissions, and deaths have been shown to occur more frequently with increasing age (6, 9, 14). In our study, age ≥65 years was the strongest independent predictor of ICU admission and in-hospital mortality. Persons aged 75–84 years had the highest the risk of ICU admission compared to 18–49 years old, and those ≥85 years experienced 11 times the risk of death. These findings are similar to other studies from China, Europe, and the United States (4, 9, 14–17). Our data provide support that older persons are particularly vulnerable to severe COVID-19 disease and should be targeted for aggressive preventive measures (8).

Being male was associated with a higher risk of ICU admission and death after adjusting for age, race/ethnicity and underlying conditions. Other studies have similarly shown male sex to be associated with COVID-19-associated hospitalizations (4, 18), ICU admissions (19), and need for mechanical ventilation (20).

Similar to other U.S. studies, we found that nearly all hospitalized patients with COVID-19 had at least one underlying medical condition (4). In contrast, underlying medical conditions were documented in only 25–50% of hospitalized cases from China (3, 21). Our analysis further demonstrated that a higher number of underlying medical conditions increased the risk of ICU admission (1.3 times the risk in persons with ≥3 vs. no underlying condition) and in-hospital mortality (1.8 times the risk in persons with ≥3 vs. no underlying condition).

In a retrospective case study among 1590 laboratory-confirmed hospitalized COVID-19 cases in 575 Chinese hospitals, Guan et al. found that after adjusting for age and smoking status, chronic obstructive pulmonary disease, diabetes, hypertension and malignancy were risk factors for a composite endpoint of ICU admission, invasive mechanical ventilation, and death (3). Similarly, we found an association between underlying medical conditions and severe outcomes, with diabetes, CLD, CVD, neurologic disease, renal disease and immunosuppression associated with in-hospital death, and diabetes, obesity, and immunosuppression associated with ICU admission. While hypertension was highly prevalent in our patient population, it was not associated with ICU admission or death. Additional studies are needed to determine whether hypertension, which is also highly prevalent in the U.S. population, increases the risk of COVID-19-associated hospitalizations and whether the duration of hypertension and the degree to which it is controlled impact the risk for severe COVID-19 disease. Similarly, the associations between the duration and degree of glycemic control in diabetes and severity of COVID-19 disease require further investigation. Obesity, which was also highly prevalent in this cohort, imparted increased risk for ICU admission, but not death. This finding may, in part, be explained by a trend of decreasing obesity prevalence with increasing age, which was a strong risk factor for mortality. Healthcare providers should be aware of these findings to appropriately triage and manage patients with high-risk conditions that may either increase risk for hospitalization or poorer outcomes once hospitalized (22, 23).

We collected data on initial symptoms, vital signs and laboratory values to characterize disease severity at admission. While approximately 70% of patients had shortness of breath at admission, the median oxygen saturation at admission was 94% on room air. Other admission vital signs and laboratory values were also largely within normal ranges. Because we did not collect data on vital signs or laboratory values during the hospital course, we may not have fully captured the onset of clinical deterioration that has been reported during the second week after illness onset (24). We limited our analysis to patients that had either been discharged or died in-hospital and found that 15% of patients received vasopressor support, and 19% received invasive mechanical ventilation. Other U.S. studies have found that up to 32% of hospitalized patients have received vasopressors and 29–33% have received invasive mechanical ventilation (19, 20), though some of these studies included patients who were still hospitalized at the time of analysis. In our study, 53% of patients requiring mechanical ventilation died, which is higher than the 36% reported in a recent study from New York City (4). In general, the proportion of patients with severe outcomes was higher in the United States than reported from China (6). Differences between patients’ outcomes in the United States and China may reflect differences in clinical practices or varying thresholds for hospitalization (18). These proportions of severe outcomes among U.S. patients are also generally higher than those found in U.S. adults hospitalized with seasonal influenza (25, 26). Our findings may help to inform resource planning and allocation in healthcare facilities during the COVID-19 pandemic.

There are several limitations to our analysis. First, it is likely that not all COVID-19-associated hospitalizations were captured because of the lack of widespread testing capability during the study period and because identification of COVID-19 patients was largely reliant on clinician-directed testing. Second, clinical practices and availability of specific interventions may differ across hospitals, which might have influenced findings. Third, COVID-NET is an ongoing surveillance system, and only 15% of the 16,318 COVID-19 hospitalized patients were included, representing those who were discharged or died in-hospital during March 1–May 2, 2020 and for whom medical records were available and chart abstractions were completed. These restrictions may have resulted in selection bias. However, there was no difference in the age and sex distribution between cases included and excluded from the analysis. The geographic distribution of cases included versus excluded from this analysis differed, which may have impacted the racial and ethnic distribution of cases included in this analysis as compared to the racial and ethnic distribution of the surveillance catchment population; however, as we do not yet have complete data on race/ethnicity for all identified cases, we were not able to assess this further. Nevertheless, COVID-NET encompasses a large geographic area with multiple hospitals and likely offers a more racially and ethnically diverse patient population compared to other single-center or state-based studies. Lastly, small counts limited our ability to determine risk factors for severe outcomes among all racial and ethnic groups. COVID-NET data will become more robust as additional medical chart reviews are completed and may allow further investigation within these racial and ethnic groups over time.

Based on preliminary findings from this multi-site, geographically diverse study, a high proportion of patients hospitalized with COVID-19 received aggressive interventions and had poor outcomes. Increasing age was the strongest predictor of in-hospital mortality. Prevention strategies, such as social distancing and rigorous hand hygiene, are key to minimizing the risk of infection in high-risk patients. These data help to characterize persons at highest risk for severe COVID-19-associated disease in the United States and to define target groups for future prevention and treatment strategies as they become available.

## Data Availability

Aggregate data is available on CDC’s COVID-NET Interactive website.

https://gis.cdc.gov/grasp/COVIDNet/COVID19_3.html

https://gis.cdc.gov/grasp/COVIDNet/COVID19_5.html

## ACKNOWLEDGMENTS

Ashley Coates, Pam Daily Kirley, Gretchen Rothrock, California Emerging Infections Program; Nisha Alden, Rachel Herlihy, Breanna Kawasaki, Colorado Department of Public Health and Environment; Paula Clogher, Hazal Kayalioglu, Amber Maslar, Adam Misiorski, Danyel Olson, Christina Parisi, Connecticut Emerging Infections Program; Kyle Openo, Emily Fawcett, Jeremiah Williams, Katelyn Lengacher, Georgia Emerging Infections Program; Andrew Weigel, Iowa Department of Health; Brian Bachaus, Timothy Blood, David Blythe, Alicia Brooks, Judie Hyun, Elisabeth Vaeth, Cindy Zerrlaut, Maryland Department of Health; Jim Collins, Kimberly Fox, Sam Hawkins, Justin Henderson, Shannon Johnson, Libby Reeg, Michigan Department of Health and Human Services; Austin Bell, Kayla Bilski, Erica Bye, Emma Contestabile, Richard Danila, Kristen Ehresmann, Hannah Friedlander, Claire Henrichsen, Emily Holodnick, Ruth Lynfield, Katherine Schliess, Samantha Siebman, Kirk Smith, Maureen Sullivan, Minnesota Department of Health; Cory Cline, New Mexico Department of Health; Kathy Angeles, Lisa Butler, Emily Hancock, Sarah Khanlian, Meaghan Novi, Sarah Shrum, New Mexico Emerging Infections Program Albuquerque; Nancy Spina, Grant Barney, Suzanne McGuire, New York State Health Department; Sophrena Bushey, Christina Felsen, Maria Gaitan, Anita Gellert, RaeAnne Kurtz, Christine Long, Shantel Peters, Marissa Tracy, University of Rochester School of Medicine and Dentistry; Laurie Billing, Maya Scullin, Jessica Shiltz, Ohio Department of Health; Nicole West, Oregon Health Authority; Kathy Billings, Katie Dyer, Anise Elie, Karen Leib, Tiffanie Markus, Terri McMinn, Danielle Ndi, Manideepthi Pemmaraju, Vanderbilt University Medical Center; Keegan McCaffrey, Utah Department of Health; Clarissa Aquino, Ryan Chatelain, Andrea George, Jacob Ortega, Andrea Price, Ilene Risk, Melanie Spencer, Ashley Swain, Salt Lake County Health Department; Mimi Huynh, Monica Schroeder, Council of State and Territorial Epidemiologists; Shua J. Chai, Field Services Branch, Division of State and Local Readiness, Center for Preparedness and Response, Centers for Disease Control and Prevention; Sharad Aggarwal, Lanson Broecker, Aaron Curns, Rebecca M. Dahl, Alexandra Ganim, Rainy Henry, Sang Kang, Sonja Nti-Berko, Robert Pinner, Mila Prill, Scott Santibanez, Alvin Shultz, Sheng-Te Tsai, Henry Walke, Venkata Akesh R. Vundi, CDC.

## Disclaimer

The conclusions, findings, and opinions expressed by the authors do not necessarily reflect the official position of the United States Department of Health and Human Services, the Public Health Service, the CDC, or the authors’ affiliated institutions.

## REFERENCES

1. Johns Hopkins University & Medicine 2020;Pages https://coronavirus.jhu.edu/map.html on 5/15/2020.

2. Centers for Disease Control & Prevention 2020;Pages https://www.cdc.gov/coronavirus/2019-ncov/covid-data/covidview/index.html#outpatient on 5/15/20220 2020.

3. Guan WJ, Liang WH, Zhao Y, Liang HR, Chen ZS, Li YM, et al. Comorbidity and its impact on 1590 patients with Covid-19 in China: A Nationwide Analysis. Eur Respir J. 2020.

4. Petrilli C, Jones S, Yang J, Rajagopalan H, O’Donnell L, Chernyak Y, et al. Factors associated with hospitalization and critical illness among 4,103 patients with COVID-19 disease in New York City. 2020.

5. COVID-19 Surveillance Group. Characteristics of COVID-19 patients dying in Italy: report based on available data on March 20th, 2020. Rome, Italy: Instituto Superiore Di Sanita; 2020.

6. Guan WJ, Ni ZY, Hu Y, Liang WH, Ou CQ, He JX, et al. Clinical Characteristics of Coronavirus Disease 2019 in China. N Engl J Med. 2020.

7. Bhatraju PK, Ghassemieh BJ, Nichols M, Kim R, Jerome KR, Nalla AK, et al. Covid-19 in Critically Ill Patients in the Seattle Region – Case Series. N Engl J Med. 2020.

8. Team CC-R. Preliminary Estimates of the Prevalence of Selected Underlying Health Conditions Among Patients with Coronavirus Disease 2019 – United States, February 12-March 28, 2020. MMWR Morb Mortal Wkly Rep. 2020;69(13):382–6.

9. Team CC-R. Severe Outcomes Among Patients with Coronavirus Disease 2019 (COVID-19) –United States, February 12-March 16, 2020. MMWR Morb Mortal Wkly Rep. 2020;69(12):343–6.

10. Garg S, Kim L, Whitaker M, O’Halloran A, Cummings C, Holstein R, et al. Hospitalization Rates and Characteristics of Patients Hospitalized with Laboratory-Confirmed Coronavirus Disease 2019 – COVID-NET, 14 States, March 1–30, 2020. MMWR Morb Mortal Wkly Rep. 2020;69(15):458–64.

11. Liang K-Y, Zeger SL. Longitudinal Data Analysis Using Generalized Linear Models.

12. Hoogendoorn WE, Bongers PM, de Vet HC, Twisk JW, van Mechelen W, Bouter LM. Comparison of two different approaches for the analysis of data from a prospective cohort study: an application to work related risk factors for low back pain. Occup Environ Med. 2002;59(7):459–65.

13. Rentsch C, Kidwai-Khan F, Tate J, Park L, King Jr. J, Skanderson M, et al. Covid-19 Testing, Hospital Admission, and Intensive Care Among 2,026,227 United States Veterans Aged 54–75 Years. 2020.

14. Chen T, Wu D, Chen H, Yan W, Yang D, Chen G, et al. Clinical characteristics of 113 deceased patients with coronavirus disease 2019: retrospective study. BMJ. 2020;368:m1091.

15. Zhou F, Yu T, Du R, Fan G, Liu Y, Liu Z, et al. Clinical course and risk factors for mortality of adult inpatients with COVID-19 in Wuhan, China: a retrospective cohort study. Lancet. 2020;395(10229):1054–62.

16. Onder G, Rezza G, Brusaferro S. Case-Fatality Rate and Characteristics of Patients Dying in Relation to COVID-19 in Italy. JAMA. 2020.

17. Wu C, Chen X, Cai Y, Xia Ja, Zhou X, Xu S, et al. Risk Factors Associated With Acute Respiratory Distress Syndrome and Death in Patients With Coronavirus Disease 2019 Pneumonia in Wuhan, China. JAMA Internal Medicine. 2020.

18. Richardson S, Hirsch JS, Narasimhan M, Crawford JM, McGinn T, Davidson KW, et al. Presenting Characteristics, Comorbidities, and Outcomes Among 5700 Patients Hospitalized With COVID-19 in the New York City Area. JAMA. 2020.

19. Myers LC, Parodi SM, Escobar GJ, Liu VX. Characteristics of Hospitalized Adults With COVID-19 in an Integrated Health Care System in California. JAMA. 2020.

20. Goyal P, Choi JJ, Pinheiro LC, Schenck EJ, Chen R, Jabri A, et al. Clinical Characteristics of Covid-19 in New York City. N Engl J Med. 2020.

21. Chen N, Zhou M, Dong X, Qu J, Gong F, Han Y, et al. Epidemiological and clinical characteristics of 99 cases of 2019 novel coronavirus pneumonia in Wuhan, China: a descriptive study. Lancet. 2020;395(10223):507–13.

22. Centers for Disease Control & Prevention 2020;Pageshttps://www.cdc.gov/coronavirus/2019-ncov/hcp/underlying-conditions.html.

23. Centers for Disease Control & Prevention 2020;Pageshttps://www.cdc.gov/coronavirus/2019-ncov/need-extra-precautions/groups-at-higher-risk.html.

24. Kujawski SA, Wong KK, Collins JP, Epstein L, Killerby ME, Midgley CM, et al. Clinical and virologic characteristics of the first 12 patients with coronavirus disease 2019 (COVID-19) in the United States. Nature Medicine. 2020.

25. Centers for Disease Control & Prevention 2020;Pageshttps://gis.cdc.gov/grasp/fluview/FluHospChars.html

26. Arriola C, Garg S, Anderson EJ, Ryan PA, George A, Zansky SM, et al. Influenza Vaccination 440 Modifies Disease Severity Among Community-dwelling Adults Hospitalized With Influenza. Clin 441 Infect Dis. 2017;65(8):1289–97.

